# Brain Structural Differences in Adults Reporting Localized Chronic Pains Mediate Risk for Suicidal Behaviors

**DOI:** 10.1101/2022.10.05.22280713

**Authors:** Ravi R. Bhatt, Elizabeth Haddad, Alyssa H. Zhu, Paul M. Thompson, Arpana Gupta, Emeran A. Mayer, Neda Jahanshad

**Affiliations:** Imaging Genetics Center, Mark and Mary Stevens Neuroimaging and Informatics Institute, Keck School of Medicine at USC, University of Southern California, Los Angeles, CA, USA; G. Oppenheimer Center for Neurobiology of Stress and Resilience, Vatche and Tamar Manoukian Division of Digestive Diseases, David Geffen School of Medicine at University of California Los Angeles, Los Angeles, CA, USA

## Abstract

**Background:** Chronic pain is a global health priority. Mapping pain occurring at different body sites, and variability in brain circuitry related to widespread chronic pain, can elucidate nuanced roles of the central nervous system underlying chronic pain conditions. Chronic pain triples suicide risk; however, whether brain circuitry can inform this risk relationship has not been investigated.

**Methods:** 11,298 participants (mean age: 64 years (range: 58-70), 55% female) with brain MRI from the UK Biobank with pain for more than 3 months in the head, neck/shoulders, back, abdomen, or hips and knees, were age-and-sex-matched to 11,298 pain-free controls. Regression models assessed cortical and subcortical structure differences between individuals reporting chronic pain and those without; mediation models determined the relationship between pain, brain structure and history of attempted suicide.

**Outcomes:** Chronic pain, regardless of site was associated with, lower surface area throughout the cortex, lower volume in the brainstem, ventral diencephalon, cerebellum, and pallidum, lower cortical thickness in the anterior insula, and greater cortical thickness in the superior parietal cortex. When differentiated by pain site, participants with chronic headaches distinctly showed an overall thicker cortex compared with controls. Chronic pain was associated with an elevated risk for suicide attempt and this relationship was mediated by lower cerebellum volume.

**Interpretation:** There are shared cortical mechanisms underlying chronic pain across body sites. An extensive thicker cortex in chronic headache was consistent with previous research. Cerebellum volume mediates the relationship between chronic pain and suicide attempt, serving as a potential biomarker prognostic for suicidal behaviors in chronic pain patients.

**Funding:** National Science Foundation, National Institutes of Health

**Research in Context:** *Evidence before this study:* Chronic pain is the leading cause of disability and disease burden globally, and its prevalence is increasing. As perception of pain occurs in the brain, alterations in brain structure have been investigated in various chronic pain conditions. However, published works, to date, report inconsistent findings, and typically do not compare a wide range of chronic pain types within the same study. Chronic pain is a risk factor for suicidal ideation, which can occur in up to 41% of individuals with chronic pain, but the role of specific brain systems in mediating the relationship between chronic pain and suicide has not been investigated.

*Added value of this study:* The present study reports alterations of brain structure in the largest and most well-powered sample reporting chronic pain to date (N = 11,298) compared to 11,298 pain-free controls, while taking into account age, sex, socioeconomic status, anxiety and depression. The effect of chronic pain on the brain is also evaluated as a function of pain across one or more of six different sites in the body (i.e. headaches, neck and shoulder, back, abdominal, hip and knee pain). Lower cortical surface area throughout the brain was related to chronic pain, and shown to be far more extensive than previously recognized. We, for the first time, show that participants with chronic headaches compared to controls have, on average, thicker gray matter throughout the cortex, a distinct and opposite pattern of effects than when individuals with other systemic pain conditions are compared to controls. A higher prevalence of suicide attempt history was noted in participants reporting chronic pain than controls. The relationship between chronic pain and suicide attempt, was mediated by the volume of the cerebellum, implicating spinocerebellar mechanisms.

*Implications of all the available evidence:* Brain structure plays a key role in chronic pain, and mediates the role between pain and suicidal behaviors, independent of commonly presenting comorbidities. Our results highlight the concept of central sensitization and the role of the brain’s interacting networks in the presence of chronic pain. The thicker cortical gray matter in chronic headaches vs. controls - compared to other chronic pain conditions - indicates different mechanisms underlie these conditions and suggests that a clinically different approach to treatment is warranted. The cerebellum volume is a reliable mediator between chronic pain and suicide attempt, a finding that provides insight into potential underlying spinocerebellar mechanisms and to how treatments such as ketamine infusions may be beneficial in chronic pain and suicidal risk behavior management. Our work shows reliable neurobiological support for the multiple brain networks impacted and in regulating mood in the chronic pain phenotype.

## Introduction

Chronic pain (CP) is a debilitating condition affecting one in five people in the United States^1^ and is the leading cause of disability and disease burden globally.^2^ It is increasingly recognized that CP is a disease of the central nervous system,^3^ and is often comorbid with psychopathology such as anxiety and depression.^4^ The theory of central sensitization underlies the central nervous system’s role in the amplification of normal sensory or mild nociceptive stimuli to produce an overwhelming and sustained pain experience, and highlights the key common mechanism underlying many distinct chronic overlapping pain conditions.^5^ Currently, measures for pain include self-report, altered behavior (e.g., avoidance and grimacing), or changes in physiology (e.g., heart or respiration rate).^6^ Neuroimaging may provide an objective measure with mechanistic insight to help decode this experience ultimately emerging in the brain.^6^ The theory behind pain processing in the brain has changed from an idea of a “pain neuromatrix”, which restricts the brain signature to pain sensation alone, to a brain signature that also considers various intrinsic brain networks and modulatory control systems that, together, produce the experience and maintenance of CP.^3,5,7^ However, neuroimaging biomarkers are still unavailable.^8^

CP is associated with an elevated risk for suicidal thoughts and behaviors.^9,10^ The World Health Organization reports that approximately 785,000 deaths by suicide occur annually, but these make up only 5% of suicide attempts.^11^ Up to 41% of individuals reporting CP also report suicidal ideation.^16^ Migraines, arthritis, back pain, and idiopathic pain, have each been independently associated with risk for suicidal behaviors, albeit to different degrees, even after controlling for age, sex, depression, and other coexisting conditions.^14,17,18^ A recent meta-analysis showed that people with CP had more prevalent death wishes, suicidal ideations, intentions, attempts, and deaths compared to people without pain (all OR > 2), with many pain-related factors such as poor sleep, poor mental health and comorbid CP conditions contributing to the elevated risk for suicide.^12^ Charting the variability in the structure and function of the brain, especially in regions involved in the perception and emotional feeling of pain, may provide mechanistic insight into how physical pain may also be associated with increased suicidal behaviors.

Here, we aim to address these gaps and inconsistencies in the literature by using data from the UK Biobank (UKB) to identify *in vivo* brain signatures of CP at large, along with localized CP subtypes, and assess the relationship of the CP brain circuitry to suicidal behaviors. The UKB provides brain MRI data from thousands of participants whose scans are collected with consistent neuroimaging protocols.^13^ This allows us to assess comorbidity across CP phenotypes, as well as CP occurring in isolation at one body site. By analyzing structural brain imaging data, we aimed to determine how brain morphometry in individuals reporting CP at different sites in the body differs relative to those not reporting CP. We further aimed to determine if these differences mediated the relationship between CP and suicidal tendencies.

## Methods

### Participants

Participant data were analyzed from the population-based UK Biobank study^13,14^ through application number 11559. Inclusion criteria included a T1-weighted brain MRI scan at the first imaging visit and a response on the pain question field ID 6159. Exclusion criteria excluded any participants diagnosed with severe mental health conditions, personality disorders, or severe intellectual disabilities, neoplasms, obesity, and diabetes, defined via ICD10 codes. Exclusionary criteria were selected to focus on potential supraspinal nociplastic mechanisms and remove confounding sources of variance.^5^ Participant demographics are summarized in **Table 1**.

### Neuroimaging Acquisition

All participants completed a 31-minute neuroimaging protocol using a Siemens Skyra 3 tesla scanner and a 32-channel head coil in one of three image scanning locations. All structural T1-weighted scans were acquired using the following parameters: 3D MPRAGE, sagittal orientation, in-plane acceleration factor = 2, TI/TR = 880/2000 ms, voxel resolution = 1 × 1 × 1 mm, acquisition matrix = 208 × 256 × 256 mm. All scans were pre-scan normalized using an on-scanner bias correction filter. More details of the imaging protocols may be found in the following.^13,14^

### Neuroimage Data Processing

Measures of regional cortical thickness (CT), regional cortical surface area (SA), and subcortical volume (VOL) were extracted using FreeSurfer 7.1.^15^ This yielded CT, SA, and VOL measurements for each of the 74 bilateral regions in the Destrieux^16^ cortical, and 11 bilateral regions plus the brainstem of the Harvard-Oxford subcortical^17–19^ atlases.

### Operational Definitions of Pain and Suicidality Variables of Interest

Participants in the UKB were asked at the time of scan about “pain types experienced in the last month” (field ID 6159), with possible answers being “Prefer not to answer”, pain at each of seven different body sites (head, face, neck/shoulder, back, stomach/abdomen, hip, knee), “all over the body”, or “None of the above”. They were then asked if the pain at the specific site had been present for three or more months (Category ID 100048). Here, CP was defined as having three or more months of pain at *any* of these body sites. Controls were defined as those who specifically reported “None of the above’’ in response to field 6159. Participants were classified as having pain at each body site if they presented with three or more months of pain at the said site, even if CP was reported at other body sites. Participants having three or more months at *exclusively* one of the body sites were classified as having exclusive CP at that body site. Participants reporting pain, but not for three months were not included in this analysis.

The presence of suicide attempt was assessed either from hospital inpatient records from intentional self-poisoning/self-harm (Category 20002, ICD-10 code X60-X84, ICD-9 E950-E959) or self-report (Category 146) of “*ever attempted suicide*” and “*attempted suicide in the past yea*r.”

### Statistical Analyses

To assess if the presence of CP was associated with cortical features, baseline neuroimaging data was compared between participants reporting CP and controls, covarying for age, sex, socioeconomic status (SES) using the Townsend deprivation index (TDI; data field 189), intracranial volume (ICV; for surface area and subcortical volume, but not cortical thickness), anxiety and depression. Participants in the CP versus healthy controls analysis were matched at a 1:1 ratio, and a 1:3 ratio for localized pain conditions, also matching for age and sex. FreeSurfer metrics of brain regions were averaged across hemispheres and subsequent analyses were computed using individual, lateralized regions (appendix). Further analyses divided up the pain conditions into those reporting chronic headaches, neck/shoulder pain, low back pain, abdominal pain, hip pain, knee pain, and pain-free controls. These participants were matched to controls with a 3:1 control-case ratio, accounting for age and sex. General linear models tested associations between each CP location and 478 brain features. Subsequent models were run adjusting for self-reported anxiety and depression. The false discovery rate (FDR) was used for multiple comparisons correction for each pain type (i.e., across all CT, SA and subcortical VOL).

To assess whether people with CP reported greater suicidal tendencies than controls, a chi-squared test was conducted between CP (yes/no) and history of suicidal attempt (yes/no). To determine if cortical structure mediates a relationship between CP and suicidal behavior, logistic regression mediations were run with CP as the predictor variable, suicide attempt as the outcome variable, and individual FDR-significant cortical and subcortical features from the case/control analyses as the mediators. Age, sex, SES, ICV, anxiety and depression were all included as covariates. All potential biological mediators were tested simultaneously with 5,000 permutations using the MultiMed package in R.^20–22^

## Results

The prevalence of CP and overlap across pain locations in the UK Biobank is depicted in **Supplementary Figure 1 and https://brainescience.shinyapps.io/ChronicPain_UKBiobank/**. Pain conditions were divided into those reporting chronic headaches (N = 1,649, mean age = 61 years, 1,157 female), neck/shoulder pain (N = 3,516, mean age = 64 years, 1,995 female), abdominal pain (N = 748, mean age= 63 years, 453 female), back pain (N = 3,891, mean age=65 years, 2,075 female), hip pain (N = 2,476, mean age = 65 years, 1,557 female), and knee pain (N = 4,628, mean age = 65 years, 2435 female). There were 26 individuals with a history of suicide attempt from hospital inpatient records, 274 individuals stating they had “*ever attempted suicide*” and 5 individuals reporting “*attempted suicide in the past year*”. In total, 291 participants had a history of at least one suicide attempt.

### Chronic Pain vs. Pain-Free, Healthy Controls

People reporting CP, irrespective of pain site, had differences in surface area, cortical thickness, and subcortical volume measures throughout the brain compared to controls (**Fig 1, Table S5**). The observed effects were not restricted to any network, but were global throughout the brain. All but eight brain regions throughout the cortex exhibited a significantly *lower* cortical surface area, the cerebellum and ventral diencephalon exhibited a *lower* volume, and the anterior insula and occipital pole exhibited a *lower* cortical thickness. Regions in the superior parietal cortex exhibited a *greater* cortical thickness.

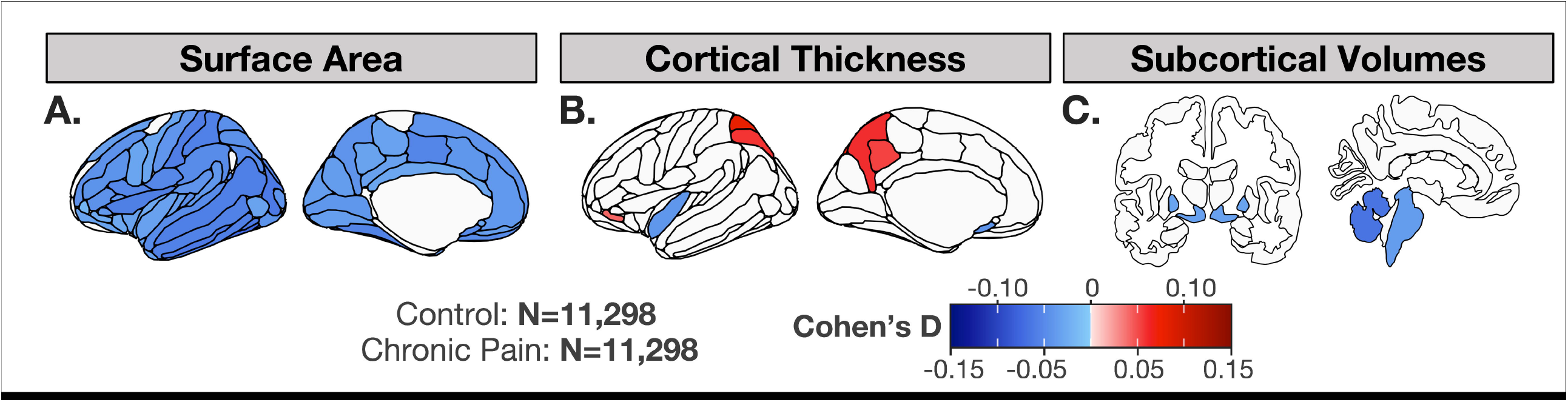

### Chronic Headaches vs. Controls

The most extensive differences in the chronic headaches subgroup were observed with cortical thickness. On average, greater cortical thickness was observed throughout the parietal cortex, occipital cortex, middle and superior frontal gyri, dorsolateral prefrontal cortex, and inferior orbital gyrus (**Fig 2, Table S5**).

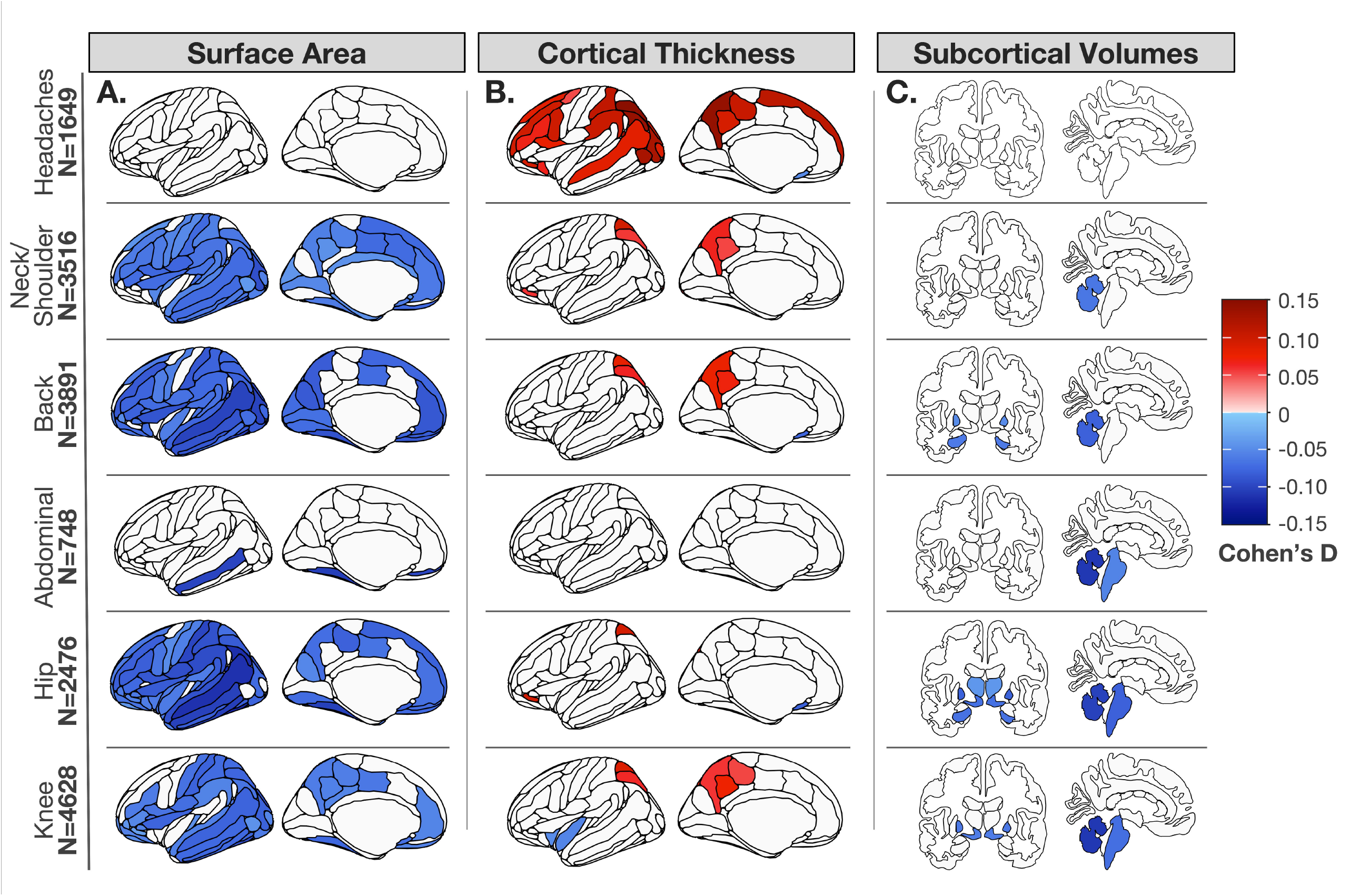

The chronic headache group showed a strikingly different pattern of surface area and cortical thickness compared to the other pain groups - a greater cortical thickness across the cortex and a lack of differences in surface area. As a post-hoc analysis, to assess if headaches showed significantly different cortical patterns compared to other pain groups, participants reporting headaches exclusively, were compared to participants reporting exclusive neck/shoulder, back, abdominal, hip and knee pain using the same procedures described above. Exploratory analyses showed a trend for greater cortical thickness in the exclusive chronic headache group (**Supplementary Material**).

### Chronic Neck/Shoulder Pain vs. Controls

People reporting chronic neck/shoulder pain had lower surface area and throughout the brain, as well as lower cerebellar volume. They also had greater cortical thickness in superior parietal regions, and the orbitofrontal cortex (**Fig 2, Table S5**).

### Chronic Back Pain vs. Controls

People reporting chronic back pain had lower surface area throughout the cortex (**Fig 2, Table S5**). Greater cortical thickness was observed in superior parietal regions, but lower in the subcallosal gyrus. Lower volumes were observed in the cerebellum, pallidum, and hippocampus.

### Chronic Abdominal Pain vs. Controls

People reporting chronic abdominal pain had on average, lower surface area in the middle temporal gyrus, fusiform face area, lingual gyrus, and suborbital sulcus. Lower volumes were observed in the cerebellum and brainstem (**Fig 2, Table S5**).

### Chronic Hip Pain vs. Controls

People reporting chronic hip pain had on average, lower surface area throughout the majority of the cortex. Greater cortical thickness was observed in the inferior orbital gyrus and superior parietal gyrus. Lower cortical thickness was observed in the subcallosal gyrus. Lower volumes were observed in the cerebellum, brainstem, pallidum, ventral diencephalon, hippocampus, and thalamus (**Fig 2, Table S5**).

### Chronic Knee Pain vs. Controls

People reporting chronic knee pain had on average, lower surface area across the occipital, temporal and parietal regions. They also had lower surface area in the dlPFC, vmPFC and OFC. They showed greater cortical thickness in the posterior parietal cortex and lower cortical thickness in the anterior insula. They showed lower subcortical volume in the cerebellum, brain stem, ventral diencephalon, and pallidum (**Table S5, Fig 2**).

Many individual participants who reported chronic pain at more than one site fell into multiple pain groups. Analyses were also performed on the subset of individuals reporting exclusively single site CP. While fewer regions were significant after FDR-correction, the distribution and direction of effects were largely in line with those of the larger group (see **Supplemental Material**).

### Post-Hoc Analyses: Chronic Headaches vs. Other Pain Conditions

People reporting chronic headaches had greater surface area vs. neck/shoulder pain, back pain, and knee pain in the sensorimotor networks, anterior insula, posterior insula, dlPFC, precuneus, temporal lobe, and occipital lobe. Greater cortical thickness was observed across occipital lobe, temporal lobe, and postcentral sulcus, as well as lower cortical thickness in the anterior cingulate compared to chronic back and knee pain. Greater volume of the cerebellum was observed in those with chronic headaches compared to abdominal and knee pain. People with chronic headaches also had significantly greater volume in the brainstem, pallidum and ventral diencephalon compared to those with chronic knee pain. All results are available in **Supplementary Tables 6-7**.

### Chronic Pain is Associated with History of Suicide Attempt

The chi-squared test revealed a significant association between suicide attempt and chronic pain (χ(1)^2^ = 20.44, *p* = 6.14 × 10^−6^, Cramer’s V = 0.03), chronic neck/shoulder pain (χ(1)^2^ = 22.74, *p* = 1.85 × 10^−6^, Cramer’s V = 0.03), chronic back pain (χ_(1)_^2^ = 18.96, *p* = 1.34 × 10^−5^, Cramer’s V = 0.03), chronic abdominal pain (χ_(1)_ ^2^= 36.15, *p* = 1.83 × 10^−9^, Cramer’s V = 0.04), chronic hip pain (χ_(1)_ ^2^= 7.89, *p* = 5.55 × 10^−3^, Cramer’s V = 0.02), and chronic knee pain (χ _(1)_ ^2^= 18.52, *p* = 1.68 × 10^−5^, Cramer’s V = 0.03). No effects were found for chronic headaches (χ_(1)_ ^2^ = 2.84, *p* = 0.09, Cramer’s V = 0.01). See **Supplemental Material** for figures and statistical details.

### Brain Structure Mediates the Relationship between Chronic Pain and History of Suicide Attempt

Cortical features determined to be significantly different between CP and controls did not mediate the relationship between chronic pain and suicide attempt. However, the effect of CP on suicidal attempt was, however, mediated via the volume of the cerebellum. A lateralized approach found the same effect on the left and right cerebellum (**Table 2**).

### Brain Structure Mediates the Relationship between Localized Chronic Pain and Suicide Attempt

The volume of the cerebellar cortex mediated the relationship between chronic neck/shoulder pain and suicide attempt. When using a lateralized analysis approach, the cortical thickness of the right superior parietal gyrus mediated the relationship between chronic neck and shoulder pain and suicide attempt. The right cerebellar cortex volume also mediated the relationship between chronic neck and shoulder pain and suicide attempt (**Table 2**).

Volume of the cerebellar cortex mediated the between chronic back pain and suicide attempt (**Table 2**). When using a lateralized analysis approach, the cortical thickness of the left precuneus and the left and right cerebellum volume also mediated the relationship between chronic back pain and suicide (**Table 2**).

The effect of chronic abdominal pain on suicide attempt was mediated via the volume of the cerebellum **(Table 2)**. A lateralized approach showed that the left cerebellar cortex mediated the relationship between CP and suicide attempt (**Table 2**).

The effect of chronic knee pain on suicide attempt was also mediated via the volume of the cerebellum. A lateralized approach showed that this was more specifically mediated by the volume of the left cerebellar cortex (**Table 2**).

## Discussion

In this work, we use *in vivo* brain MRI data from 11,298 adults reporting CP, and 11,298 matched controls to advance our understanding the role of the CNS in pain in three ways; we set out to (1) identify differences in brain morphometry between individuals with CP, and controls; (2) identify the overlap in brain systems across localized pain types, and identify *post hoc* the distinct signature of headaches; (3) confirm the relationship between CP and suicide attempt, while identifying how variation in certain brain structures, including cerebellar volume, may mediate this relationship.

Extensive differences regarding cortical surface area, thickness, and subcortical volumes were observed in all CP conditions versus controls. Gray matter differences between people with CP vs controls had previously shown *lower* gray matter volume throughout the brain, including the prefrontal cortex,^23^ insula, thalamus, medial temporal lobe, the anterior and posterior cingulate, hippocampal regions, basal ganglia, sensorimotor network, brainstem, cerebellum and ventral diencephalon. However, *larger* gray matter volumes have also been observed.^3,24–31^ Our highly-powered results break volume down into the biologically divergent thickness and surface area components, helping to explain the sources of discrepancies in the literature. Using a much larger sample size, we also show cortical and subcortical gray matter differences associated with CP are more extensive than previously reported. The effects are still present even when controlling for anxiety and depression. This highlights the need for large, highly powered studies for accurate and replicable results, with the potential for use in clinical settings.^32^

Chronic back, neck and shoulder, hip and knee pain showed similar results compared to the full sample in terms of cortical surface area and volume, albeit not as extensive. The hippocampus was consistently involved, highlighting the emerging mechanistic role the hippocampus and its extracellular matrix^33–36^ plays in CP. The pallidum (i.e., globus pallidus) was also consistently significant across pain groups. The role of the basal ganglia in sensorimotor integration in response to pain and noxious stimuli is well established.^37–39^ Deep brain stimulation of the globus pallidus has been shown to relieve sensory deficits, and improve and maintain pain symptoms over time.^40,41^

People with chronic headaches exhibited a largely different pattern of brain differences than was found in the other CP conditions, as compared to controls, specifically thicker cortical gray matter throughout the cortex. Individuals with exclusive chronic headaches showed a significantly greater cortical thickness in the occipital pole than pain-free controls. Greater cortical thickness has been observed in other studies in people reporting chronic headaches and migraine, mainly located in the primary somatosensory cortex, temporo-occipital and middle frontal regions,^42–47^ however no study has reported higher cortical thickness throughout the cortex to the extent identified in this study. Cortical spreading depression (CSD) is the current leading theory underlying the pathophysiology of migraine.^46,48,49^ CSD is characterized as a neuronal and glial depolarization originating at the occipital pole and spreading anteriorly over the lateral, medial and ventral surfaces of the brain.^46,48^ A plausible theory as to why a thicker cortex would be observed across the brain where CSD occurs is that an influx of water follows the influx of sodium and calcium in neurons, and sodium and potassium pumps fail to provide a sufficient outward current to balance the inward currents of AMPA/kainate receptor and NMDA receptor channels.^46,50^ This theory needs further validation in human chronic headache/migraine populations. A recent study modeling CSD propagation in migraine patients using diffusion MRI showed the propagation occurs in regions overlapping with the thicker regions identified in this study, specifically the visual cortex, sensorimotor network, parietal lobe, Broca’s area, and the dlPFC.^51^ The thicker occipital pole in chronic exclusive headaches suggests the originating point of CSD and should be further investigated.

Moreover, CP as a whole was associated with suicide attempt, as were many of the individual CP subtypes, as defined by body site. Chronic abdominal pain had the greatest association with suicide attempt. When investigating CP as a whole, the cerebellar volume consistently mediated the relationship between CP and suicide attempt. The cerebellum receives top-down inputs from the somatosensory cortices, thalamus, limbic structures, and periaqueductal gray and bottom-up inputs from the spinal cord, it is well positioned to have an influence on pain perception and aversive emotional processing.^52^ The cerebellum is increasingly recognized for its role in negative emotion processing and similar underlying circuitry of aversive emotional processing and pain.^52^ The observed lower volume is consistent with past research in suicide attempters.^53^ A meta-analysis showed differentially methylated regions of DNA in the cerebellum of suicide completers largely independent of comorbid psychiatric disorders, which are involved in cyclin E expression, schizophrenia susceptibility, and preserving neuronal health.^54^

Improvement in suicidal ideation after open-label ketamine infusion has been linked to increases in regional glucose metabolism (rCMRglu) in the cerebellum.^55^ Improvements after ketamine infusion have been associated with changes in functional connectivity between the cerebellum and large scale functional networks in those with major depression.^56^ Engaging the descending pain modulation pathways and the cerebellum in those with chronic pain.^57^ Multiple nociceptive spinocerebellar pathways that overlap with aversive emotional processing have been proposed.^58–60^ Neuroimaging evidence suggests that overlapping regions of the cerebellum - including VI, VIIb and *crus* I - process nociceptive and aversive emotional stimuli, which are inversely correlated with activation in limbic areas.^61^ This suggests that the cerebellum is not specific to pain processing, but to aversive sensory and affective processing more generally, playing a key role in suicidal behaviors.^52^

The right superior parietal gyrus was a key region in mediating the relationship between chronic neck and shoulder pain and suicide attempt. The superior parietal gyrus plays a key role in sensorimotor integration by maintaining an internal representation of the body’s state,^62^ and is a region consistently implicated in multisensory integration of pain and altered higher level pain processes in those with CP.^3^ Neck and shoulder pain often co-occur with and can even aggravate headaches, and our sample showed the most overlap with chronic headaches and neck/shoulder pain. This can be clinically observed with tenderness, tightness, and muscle contractions in myofascial trigger points in the neck and shoulders, that leads to a “pull” of the dura mater and formation of myodural bridges.^63^ The current results may also corroborate recent functional imaging research. Functional connectivity between the superior parietal gyrus and the middle frontal gyrus – known as the executive control network^64^ – is lower in suicide attempters and was associated with greater suicidal ideation and impulsivity.^65,66^

As this investigation is cross-sectional, no conclusions can be made about causal relationships between the observed brain changes, CP, and suicide attempts. The sample sizes for the exclusive pain sites were also significantly lower compared to the non-exclusive pain sites. This decrease in power is likely a reason for the difference in the significant number of regional effects between groups. Large samples are needed when assessing brain-wide associations in neuroimaging research.^67^ Although well powered, the UK Biobank consists of data from largely middle-to older-aged British white individuals. Replicability and generalizability of these results should be assessed in people from a variety of age ranges and ethnicities. Future studies should also use other modalities in addition to gray matter morphology to better understand mechanistic reasons underlying the brain structural differences observed in the current study.

Our study reports differences in brain structure compared to controls in the largest CP sample to date, specific to pain location on the body, and how these differences mediate the relationship between CP and suicide attempt. CP undifferentiated by body site was associated with lower surface area throughout the cortex, lower volume in the brainstem, ventral diencephalon, cerebellum and pallidum, lower cortical thickness in the anterior insula, and greater cortical thickness in the superior parietal lobe. When differentiated by body site, similar patterns remained. However, the chronic headache group exhibited greater cortical thickness throughout the cortex. This follows a pattern of cortical spreading depression originating in the occipital lobe and spreading anteriorly across the brain. There was an association between CP and suicide attempt, that remained across body sites. The cerebellum consistently mediated the relationship between CP and suicide attempt, suggesting the involvement of nociceptive spinocerebellar pathways responsible for processing aversive nociceptive and affective processing, underlying the relationship between CP and suicide attempt.

## Data Availability

The individual-level MRI and phenotype UK Biobank data are available upon application to UK Biobank (https://www.ukbiobank.ac.uk/register-apply/).

https://brainescience.shinyapps.io/ChronicPain_UKBiobank/

## Notes

**Conflict of Interest:** NJ and PT were MPI of a grant from Biogen, Inc., for work unrelated to the contents of this manuscript. AG is a scientific consultant for Yamaha on works unrelated to the contents of this manuscript. EAM is a member of the scientific advisory boards of Danone, Axial Therapeutics, Amare, Mahana Therapeutics, Pendulum, Bloom Biosciences, and APC Microbiome Ireland.

**Support:** Supported by NIH grants R01 MH117601 (Jahanshad), R01 AG059874 (Jahanshad), P50 DK064539 (Mayer), U01 DK082370 (Mayer), R01 MD015904 (AG), R03 DK121025 (AG), NSF GRFP 2020290241 (Bhatt), UK Biobank Resource Application 11559.

### Competing Interest Statement

Conflict of Interest: NJ and PT were MPI of a grant from Biogen, Inc., for work unrelated to the contents of this manuscript. AG is a scientific consultant for Yamaha on works unrelated to the contents of this manuscript. EAM is a member of the scientific advisory boards of Danone, Axial Therapeutics, Amare, Mahana Therapeutics, Pendulum, Bloom Biosciences, and APC Microbiome Ireland.

### Funding Statement

This study was supported by NIH grants R01 MH117601 (Jahanshad), R01 AG059874 (Jahanshad), P50 DK064539 (Mayer), U01 DK082370 (Mayer), R01 MD015904 (AG), R03 DK121025 (AG), NSF GRFP 2020290241 (Bhatt), UK Biobank Resource Application 11559.

### Author Declarations

The individual-level MRI and phenotype UK Biobank data are available upon application to UK Biobank (https://www.ukbiobank.ac.uk/register-apply/). Data were accessed under UK Biobank Resource Application 11559.

