## Supplemental Material for "Brain Structural Differences in Adults Reporting Localized Chronic Pains Mediate Risk for Suicidal Behaviors"

**Supplement to Morphological Brain Differences Mediate Suicidal Tendencies in People Reporting Chronic Pain in the UK Biobank**

**Table of Contents**

- sFigure 1: Prevalence of chronic pain the UK Biobank
- sFigure 2: Association between chronic pain and suicide
- sFigure 3: Differences in brain structure in participants reporting chronic pain exclusively across different body regions compared to controls.
- sFigure 4: Differences in brain structure using a lateralized approach in participants reporting chronic pain across different body regions compared to controls.
- sFigure 5: Differences in brain structure using a lateralized approach in participants reporting exclusive chronic pain across different body regions compared to controls.
- sTable 1: Demographics participants with chronic pain vs. controls
- sTable 2: Demographics of participants with chronic back pain vs. controls
- sTable 3: Demographics of participants with exclusive chronic back pain vs. controls
- sTable 4: Demographics of participants with chronic headaches vs. controls.
- sTable 5: Demographics of participants with chronic exclusive headaches vs. controls
- sTable 6: Full hemisphere averaged brain results of various chronic pain conditions vs. controls
- sTable 7: Full lateralized brain results of various chronic pain conditions vs. controls.

**Methods:**

Statistical Analysis: To asses differences between chronic pain groups and controls, either wilxcon rank To assess whether people with CP reported greater suicidal tendencies than controls, a chi-squared test was conducted between CP (yes/no) and history of suicidal attempt (yes/no). To determine if cortical structure mediates the relationship between CP and suicidal tendencies, logistic regression mediations were run with the presence of chronic pain as the predictor variable, the presence of suicide attempt as the outcome variable, and FDR significant cortical and subcortical features from the case/control analyses as the mediator. Age, sex, SES, and ICV were all included as covariates. All potential biological mediators were tested simultaneously with 5000 permutations using the MultiMed package in R^1,2^. Briefly put, this is a permutation based method with joint correction based on the maximal test statistic and has 2-5 times more power than Bonferroni correction. The outcome is an S-statistic and a p-value. The S-statistic is the absolute value of the product of the correlation between the independent variable and the mediator and the partial correlation between the mediator and the outcome conditional on the independent variable. A greater S-statistic and lower p-value is indicative of a more significant mediator.^1,2^ Unstandardized indirect effects were computed for each of 5,000 bootstrapped samples, and the 95% confidence interval was computed by determining the indirect effects at the 2.5th and 97.5th percentiles using the mediation package^3^ in R.

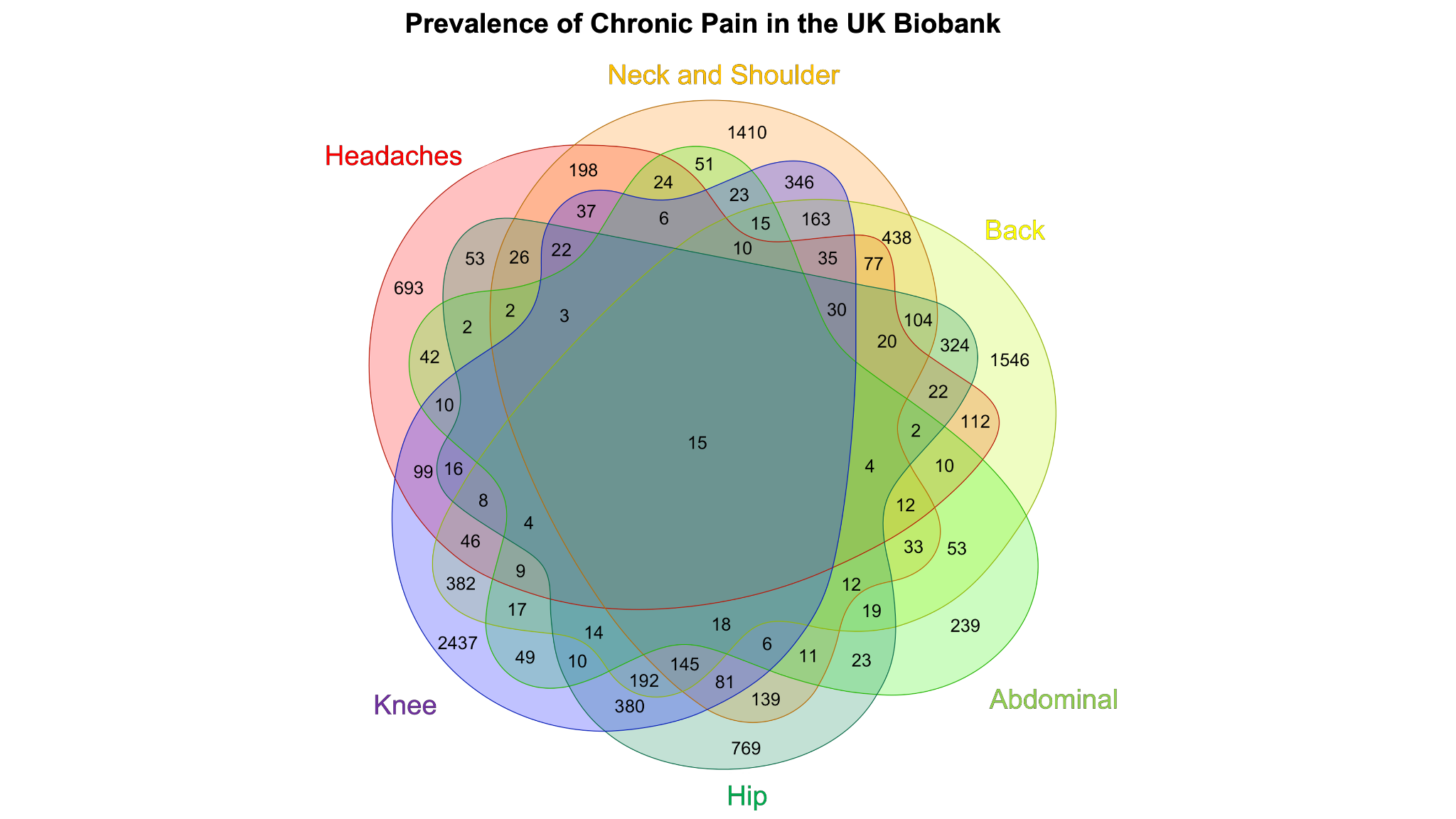

sFigure 1: Counts of chronic pain conditions by site and their overlap. Numbers represent the amount of people with each condition and their overlap with other pain conditions. Total N = 11,298.

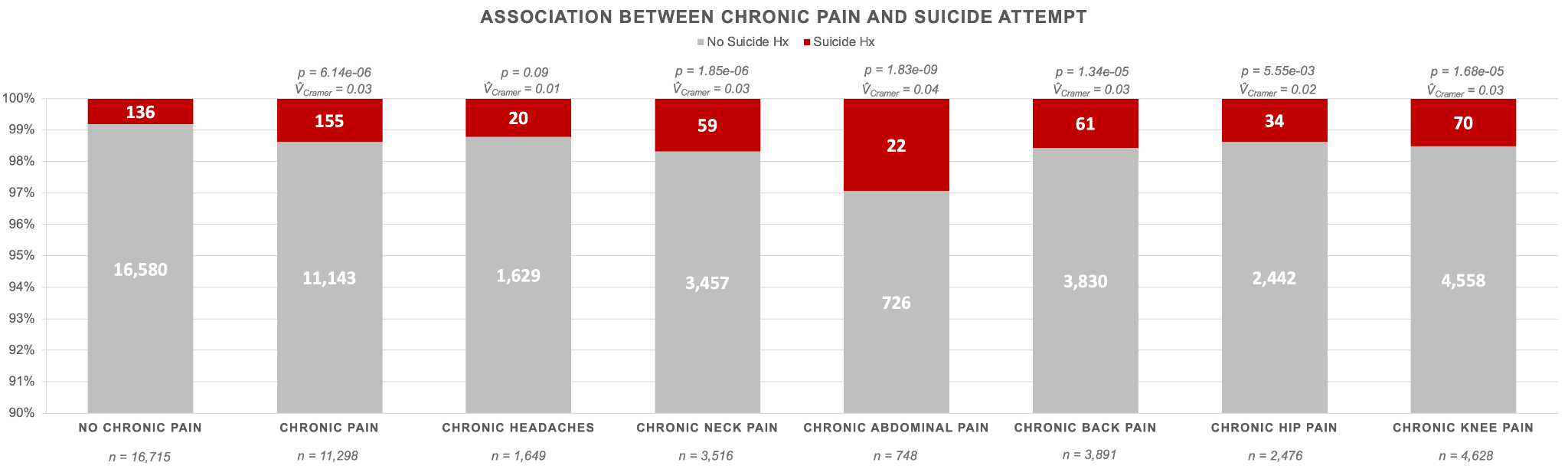

sFigure 2: Proportion of individuals reporting sucide attempt by chronic pain group. Chi-squared tests were conducted between no chronic pain and each of the chronic pain groups. Significance values and effect sizes are reported for each group. *p*: p-value, *V* = Cramer’s V

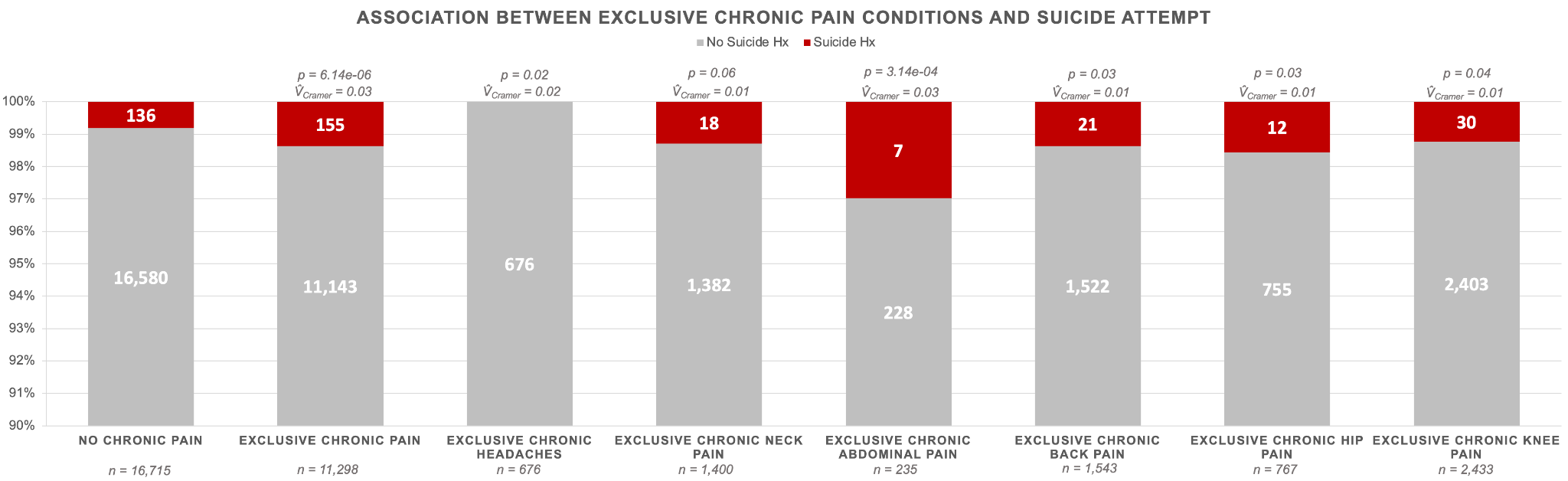

sFigure 3: Proportion of individuals reporting suicide attempt by exclusive chronic pain group. Chi-squared tests were conducted between no chronic pain and each of the exclusive chronic pain groups. Significance values and effect sizes are reported for each group. *p*: p-value, *V* = Cramer’s V

sTable 1: Demographics of participants with chronic headaches matched to controls by age and sex

| **Characteristic** | **N** | **Chronic Headaches, N = 1 649^1^** | **No Chronic Pain, N = 4 947^1^** | **p-value^2^** | **Cohen's D** | **Cramer's V** |
| --- | --- | --- | --- | --- | --- | --- |
| **Age** | 6 596 | 61 (55 – 67) | 61 (55 – 67) | 0.997 | -1.77e-04 |  |
| **Sex** | 6 596 |  |  | 1 |  | 0.00 |
| Female |  | 1 157 (70%) | 3 471 (70%) |  |  |  |
| Male |  | 492 (30%) | 1 476 (30%) |  |  |  |
| **TDI** | 4 925 | 2·58 (0·18 – 3·88) | 2·52 (0·44 – 3·87) | 0.892 | -0.324 |  |
| Unknown |  | 836 | 835 |  |  |  |
| **eTIV mm^3^** | 6 596 | 1.4e+06 (1.32e+06 – 1.5e+06) | 1.4e+06 (1.32e+06 – 1.5e+06) | 0.741 | 0.0148 |  |
| **Anxiety** | 4 813 | 540 (68%) | 1 939 (48%) | 9.30e-25 |  | 0.148 |
| Unknown |  | 857 | 926 |  |  |  |
| **Depression** | 4 837 | 272 (35%) | 579 (14%) | 8.57e-43 |  | 0.196 |
| Unknown |  | 864 | 895 |  |  |  |
| ^1^Median (IQR); n (%) | | | | | | |
| ^2^Wilcoxon rank sum test; Pearson's Chi-squared test | | | | | | |

sTable 2: Demographics of participants with chronic exclusive headaches matched to controls by age and sex

| **Characteristic** | **N** | **Chronic Exclusive Headaches, N = 676^1^** | **No Chronic Pain, N = 2 028^1^** | **p-value^2^** | **Cohen's D** | **Cramer's V** |
| --- | --- | --- | --- | --- | --- | --- |
| **Age** | 2 704 | 61 (55 – 66) | 61 (55 – 66) | 1 | -1.06e-04 |  |
| **Sex** | 2 704 |  |  | 1 |  | 0.00 |
| Female |  | 461 (68%) | 1 383 (68%) |  |  |  |
| Male |  | 215 (32%) | 645 (32%) |  |  |  |
| **TDI** | 1 545 | NA (NA – NA) | 2·55 (0·44 – 3·95) |  | 0.673 |  |
| Unknown |  | 676 | 483 |  |  |  |
| **eTIV mm^3^** | 2 704 | 1.41e+06 (1.33e+06 – 1.52e+06) | 1.40e+06 (1.32e+06 – 1.50e+06) | 0.0284 | 0.111 |  |
| **Anxiety** | 1 515 | 0 (NA%) | 744 (49%) |  |  |  |
| Unknown |  | 676 | 513 |  |  |  |
| **Depression** | 1 522 | 0 (NA%) | 243 (16%) |  |  |  |
| Unknown |  | 676 | 506 |  |  |  |
| ^1^Median (IQR); n (%) | | | | | | |
| ^2^Wilcoxon rank sum test; Pearson's Chi-squared test | | | | | | |

sTable 3: Demographics of participants with chronic neck pain matched to controls by age and sex

| **Characteristic** | **N** | **Chronic Neck Pain, N = 3 516^1^** | **No Chronic Pain, N = 10 548^1^** | **p-value^2^** | **Cohen's D** | **Cramer's V** |
| --- | --- | --- | --- | --- | --- | --- |
| **Age** | 14 064 | 64 (58 – 70) | 64 (58 – 70) | 0.998 | 4.85e-04 |  |
| **Sex** | 14 064 |  |  | 1 |  | 0.00 |
| Female |  | 1 995 (57%) | 5 985 (57%) |  |  |  |
| Male |  | 1 521 (43%) | 4 563 (43%) |  |  |  |
| **TDI** | 12 434 | 2·66 (0·42 – 3·91) | 2·63 (0·59 – 3·91) | 0.654 | -2.44e-02 |  |
| Unknown |  | 479 | 1 151 |  |  |  |
| **eTIV mm^3^** | 14 064 | 1.41e+06 (1.32e+06 – 1.52e+06) | 1.42e+06 (1.33e+06 – 1.52e+06) | 0.0467 | -1.42e-02 |  |
| **Anxiety** | 12 122 | 1 739 (59%) | 4 142 (45%) | 5.49e-37 |  | 0.115 |
| Unknown |  | 551 | 1 391 |  |  |  |
| **Depression** | 12 174 | 736 (25%) | 1 182 (13%) | 1.81e-57 |  | 0.145 |
| Unknown |  | 587 | 1 303 |  |  |  |
| ^1^Median (IQR); n (%) | | | | | | |
| ^2^Wilcoxon rank sum test; Pearson's Chi-squared test | | | | | | |

sTable 4: Demographics of participants with chronic exclusive neck pain matched to controls by age and sex

| **Characteristic** | **N** | **Chronic Exclusive Neck Pain, N = 1 400^1^** | **No Chronic Pain, N = 4 200^1^** | **p-value^2^** | **Cohen's D** | **Cramer's V** |
| --- | --- | --- | --- | --- | --- | --- |
| **Age** | 5 600 | 64 (58 – 71) | 64 (58 – 71) | 1 | 9.05e-05 |  |
| **Sex** | 5 600 |  |  | 1 |  | 0.00 |
| Female |  | 711 (51%) | 2 133 (51%) |  |  |  |
| Male |  | 689 (49%) | 2 067 (49%) |  |  |  |
| **TDI** | 4 622 | 2·78 (0·79 – 3·99) | 2·70 (0·59 – 3·94) | 0.118 | 5.62e-02 |  |
| Unknown |  | 154 | 824 |  |  |  |
| **eTIV mm^3^** | 5 600 | 1.43e+06 (1.33e+06 – 1.53e+06) | 1.43e+06 (1.34e+06 – 1.53e+06) | 0.365 | 6.76e-03 |  |
| **Anxiety** | 4 510 | 658 (54%) | 1 451 (44%) | 2.27e-09 |  | 0.0885 |
| Unknown |  | 183 | 907 |  |  |  |
| **Depression** | 4 533 | 227 (19%) | 433 (13%) | 1.31e-06 |  | 0.0712 |
| Unknown |  | 190 | 877 |  |  |  |
| ^1^Median (IQR); n (%) | | | | | | |
| ^2^Wilcoxon rank sum test; Pearson's Chi-squared test | | | | | | |

sTable 5: Demographics of participants with chronic abdominal pain matched to controls by age and sex

| **Characteristic** | **N** | **Chronic Abdominal Pain, N = 748^1^** | **No Chronic Pain, N = 2 244^1^** | **p-value^2^** | **Cohen's D** | **Cramer's V** |
| --- | --- | --- | --- | --- | --- | --- |
| **Age** | 2 992 | 63 (57 – 69) | 63 (57 – 69) | 1 | 5.52e-05 |  |
| **Sex** | 2 992 |  |  | 1 |  | 0.00 |
| Female |  | 453 (61%) | 1 359 (61%) |  |  |  |
| Male |  | 295 (39%) | 885 (39%) |  |  |  |
| **TDI** | 2 252 | 2·22 (0·00 – 3·82) | 2·52 (0·60 – 3·80) | 0.0549 | -0.110 |  |
| Unknown |  | 119 | 621 |  |  |  |
| **eTIV mm^3^** | 2 992 | 1.42e+06 (1.33e+06 – 1.52e+06) | 1.41e+06 (1.32e+06 – 1.52e+06) | 0.768 | -2.11e-03 |  |
| **Anxiety** | 2 215 | 408 (66%) | 702 (44%) | 5.29e-20 |  | 0.194 |
| Unknown |  | 127 | 650 |  |  |  |
| **Depression** | 2 213 | 202 (33%) | 209 (13%) | 4.89e-27 |  | 0.228 |
| Unknown |  | 135 | 644 |  |  |  |
| ^1^Median (IQR); n (%) | | | | | | |
| ^2^Wilcoxon rank sum test; Pearson's Chi-squared test | | | | | | |

sTable 6: Demographics of participants with chronic exclusive abdominal pain matched to controls by age and sex

| **Characteristic** | **N** | **Chronic Exclusive Abdominal Pain, N = 235^1^** | **No Chronic Pain, N = 16 716^1^** | **p-value^2^** | **Cohen's D** | **Cramer's V** |
| --- | --- | --- | --- | --- | --- | --- |
| **Age** | 16 951 | 63 (57 – 70) | 65 (59 – 70) | 0.0134 | -0.167 |  |
| **Sex** | 16 951 |  |  | 0.114 |  | 0.0116 |
| Female |  | 129 (55%) | 8 309 (50%) |  |  |  |
| Male |  | 106 (45%) | 8 407 (50%) |  |  |  |
| **TDI** | 15 544 | 2·36 (0·52 – 3·99) | 2·68 (0·62 – 3·93) | 0.458 | -5.71e-02 |  |
| Unknown |  | 23 | 1 384 |  |  |  |
| **eTIV mm^3^** | 16 951 | 1.43e+06 (1.34e+06 – 1.50e+06) | 1.43e+06 (1.34e+06 – 1.54e+06) | 0.162 | -0.119 |  |
| **Anxiety** | 15 164 | 130 (63%) | 6 566 (44%) | 5.35e-08 |  | 0.0436 |
| Unknown |  | 28 | 1 759 |  |  |  |
| **Depression** | 15 279 | 56 (27%) | 1 804 (12%) | 7.57e-11 |  | 0.0518 |
| Unknown |  | 26 | 1 646 |  |  |  |
| ^1^Median (IQR); n (%) | | | | | | |
| ^2^Wilcoxon rank sum test; Pearson's Chi-squared test | | | | | | |

sTable 7: Demographics of participants with chronic back pain matched to controls by age and sex

| **Characteristic** | **N** | **Chronic Back Pain, N = 3 891^1^** | **No Chronic Pain, N = 11 673^1^** | **p-value^2^** | **Cohen's D** | **Cramer's V** |
| --- | --- | --- | --- | --- | --- | --- |
| **Age** | 15 564 | 65 (58 – 70) | 65 (58 – 70) | 0.985 | 7.34e-04 |  |
| **Sex** | 15 564 |  |  | 1 |  | 0.00 |
| Female |  | 2 075 (53%) | 6 225 (53%) |  |  |  |
| Male |  | 1 816 (47%) | 5 448 (47%) |  |  |  |
| **TDI** | 13 757 | 2·61 (0·28 – 3·90) | 2·65 (0·60 – 3·93) | 0.0793 | -0.0478 |  |
| Unknown |  | 596 | 1 211 |  |  |  |
| **eTIV mm^3^** | 15 564 | 1.41e+06 (1.32e+06 – 1.52e+06) | 1.43e+06 (1.33e+06 – 1.53e+06) | 2.65e-05 | -0.0528 |  |
| **Anxiety** | 13 404 | 1 828 (57%) | 4 536 (45%) | 9.25e-35 |  | 0.106 |
| Unknown |  | 680 | 1 480 |  |  |  |
| **Depression** | 13 460 | 789 (25%) | 1 237 (12%) | 1.27e-68 |  | 0.151 |
| Unknown |  | 701 | 1 403 |  |  |  |
| ^1^Median (IQR); n (%) | | | | | | |
| ^2^Wilcoxon rank sum test; Pearson's Chi-squared test | | | | | | |

sTable 8: Demographics of participants with chronic exclusive back pain matched to controls by age and sex

| **Characteristic** | **N** | **Chronic Exclusive Back Pain, N = 1 543^1^** | **No Chronic Pain, N = 4 629^1^** | **p-value^2^** | **Cohen's D** | **Cramer's V** |
| --- | --- | --- | --- | --- | --- | --- |
| **Age** | 6 172 | 65 (59 – 71) | 65 (59 – 71) | 0.99 | -1.31e-04 |  |
| **Sex** | 6 172 |  |  | 1 |  | 0.00 |
| Female |  | 716 (46%) | 2 148 (46%) |  |  |  |
| Male |  | 827 (54%) | 2 481 (54%) |  |  |  |
| **TDI** | 5 077 | 2·71 (0·60 – 3·91) | 2·68 (0·60 – 3·90) | 0.898 | 0.00976 |  |
| Unknown |  | 210 | 885 |  |  |  |
| **eTIV mm^3^** | 6 172 | 1.43e+06 (1.34e+06 – 1.53e+06) | 1.44e+06 (1.34e+06 – 1.54e+06) | 0.0208 | -0.0461 |  |
| **Anxiety** | 4 945 | 662 (51%) | 1 566 (43%) | 3.89e-07 |  | 0.0717 |
| Unknown |  | 247 | 980 |  |  |  |
| **Depression** | 4 966 | 249 (19%) | 440 (12%) | 6.76e-11 |  | 0.0919 |
| Unknown |  | 251 | 955 |  |  |  |
| ^1^Median (IQR); n (%) | | | | | | |
| ^2^Wilcoxon rank sum test; Pearson's Chi-squared test | | | | | | |

sTable 9: Demographics of participants with chronic hip pain matched to controls by

age and sex

| **Characteristic** | **N** | **Chronic Hip Pain, N = 2 476^1^** | **No Chronic Pain, N = 7 428^1^** | **p-value^2^** | **Cohen's D** | **Cramer's V** |
| --- | --- | --- | --- | --- | --- | --- |
| **Age** | 9 904 | 65 (59 – 70) | 65 (59 – 70) | 0.998 | 7.70e-05 |  |
| **Sex** | 9 904 |  |  | 1 |  | 0.00 |
| Female |  | 1 557 (63%) | 4 671 (63%) |  |  |  |
| Male |  | 919 (37%) | 2 757 (37%) |  |  |  |
| **TDI** | 8 556 | 2·55 (0·30 – 3·89) | 2·69 (0·62 – 3·96) | 0.0554 | -0.0489 |  |
| Unknown |  | 344 | 1 004 |  |  |  |
| **eTIV mm^3^** | 9 904 | 1.39e+06 (1.31e+06 – 1.50e+06) | 1.41e+06 (1.32e+06 – 1.51e+06) | 0.00205 | -0.0407 |  |
| **Anxiety** | 8 342 | 1 147 (55%) | 2 896 (46%) | 9.52e-13 |  | 0.0779 |
| Unknown |  | 400 | 1 162 |  |  |  |
| **Depression** | 8 384 | 480 (23%) | 759 (12%) | 1.02e-35 |  | 0.136 |
| Unknown |  | 410 | 1 110 |  |  |  |
| ^1^Median (IQR); n (%) | | | | | | |
| ^2^Wilcoxon rank sum test; Pearson's Chi-squared test | | | | | | |

sTable 10: Demographics of participants with chronic exclusive hip pain matched to controls by age and sex

| Characteristic | N | Chronic Exclusive Hip Pain, N = 767^1^ | No Chronic Pain, N = 2 301^1^ | p-value^2^ | Cohen's D | Cramer's V |
| --- | --- | --- | --- | --- | --- | --- |
| **Age** | 3 068 | 65 (59 – 71) | 65 (59 – 71) | 0.99 | 4.35e-05 |  |
| **Sex** | 3 068 |  |  | 1 |  | 0.00 |
| Female |  | 477 (62%) | 1 431 (62%) |  |  |  |
| Male |  | 290 (38%) | 870 (38%) |  |  |  |
| **TDI** | 2 350 | 2·60 (0·79 – 3·97) | 2·63 (0·66 – 3·95) | 0.528 | 0.0476 |  |
| Unknown |  | 67 | 651 |  |  |  |
| **eTIV mm^3^** | 3 068 | 1.40e+06 (1.32e+06 – 1.51e+06) | 1.41e+06 (1.32e+06 – 1.51e+06) | 0.510 | 0.0197 |  |
| **Anxiety** | 2 289 | 340 (51%) | 754 (47%) | 0.0760 |  | 0.0361 |
| Unknown |  | 96 | 683 |  |  |  |
| **Depression** | 2 307 | 122 (18%) | 192 (12%) | 1.43e-04 |  | 0.0778 |
| Unknown |  | 81 | 680 |  |  |  |
| ^1^Median (IQR); n (%) | | | | | | |
| ^2^Wilcoxon rank sum test; Pearson's Chi-squared test | | | | | | |

sTable 11: Demographics of participants with chronic knee pain matched to controls by

age and sex

| **Characteristic** | **N** | **Chronic Knee Pain, N = 4 628^1^** | **No Chronic Pain, N = 13 884^1^** | **p-value^2^** | **Cohen's D** | **Cramer's V** |
| --- | --- | --- | --- | --- | --- | --- |
| **Age** | 18 512 | 65 (58 – 70) | 65 (59 – 70) | 0.989 | 6.84e-04 |  |
| **Sex** | 18 512 |  |  | 1 |  | 0.00 |
| Female |  | 2 435 (53%) | 7 305 (53%) |  |  |  |
| Male |  | 2 193 (47%) | 6 579 (47%) |  |  |  |
| **TDI** | 16 628 | 2·55 (0·35 – 3·84) | 2·68 (0·62 – 3·93) | 0.00329 | -0.0495 |  |
| Unknown |  | 591 | 1 293 |  |  |  |
| **eTIV mm^3^** | 18 512 | 1.42e+06 (1.32e+06 – 1.52e+06) | 1.43e+06 (1.33e+06 – 1.53e+06) | 0.00437 | -0.0403 |  |
| **Anxiety** | 16 213 | 2 091 (53%) | 5 454 (44%) | 1.19e-21 |  | 0.0749 |
| Unknown |  | 694 | 1 605 |  |  |  |
| **Depression** | 16 293 | 820 (21%) | 1 511 (12%) | 3.50e-42 |  | 0.106 |
| Unknown |  | 713 | 1 506 |  |  |  |
| ^1^Median (IQR); n (%) | | | | | | |
| ^2^Wilcoxon rank sum test; Pearson's Chi-squared test | | | | | | |

sTable 12: Demographics of participants with chronic exclusive knee pain matched to controls by age and sex

| **Characteristic** | **N** | **Chronic Exclusive Knee Pain, N = 2 433^1^** | **No Chronic Pain, N = 7 299^1^** | **p-value^2^** | **Cohen's D** | **Cramer's V** |
| --- | --- | --- | --- | --- | --- | --- |
| **Age** | 9 732 | 65 (59 – 70) | 65 (59 – 70) | 0.99 | 8.28e-05 |  |
| **Sex** | 9 732 |  |  | 1 |  | 0.00 |
| Female |  | 1 181 (49%) | 3 543 (49%) |  |  |  |
| Male |  | 1 252 (51%) | 3 756 (51%) |  |  |  |
| **TDI** | 8 462 | 2·58 (0·54 – 3·83) | 2·67 (0·61 – 3·94) | 0.116 | -0.0226 |  |
| Unknown |  | 262 | 1 008 |  |  |  |
| **eTIV mm^3^** | 9 732 | 1.43e+06 (1.33e+06 – 1.54e+06) | 1.43e+06 (1.34e+06 – 1.54e+06) | 0.550 | -0.0298 |  |
| **Anxiety** | 8 255 | 1 024 (49%) | 2 662 (43%) | 2.66e-05 |  | 0.0460 |
| Unknown |  | 325 | 1 152 |  |  |  |
| **Depression** | 8 313 | 363 (17%) | 738 (12%) | 6.45e-10 |  | 0.0674 |
| Unknown |  | 320 | 1 099 |  |  |  |
| ^1^Median (IQR); n (%) | | | | | | |
| ^2^Wilcoxon rank sum test; Pearson's Chi-squared test | | | | | | |

*Chronic Exclusive Headaches vs. Controls*

In the subset of 676 people reporting exclusive chronic headaches (no other pain site), greater cortical thickness was observed in the anterior occipital sulcus and the middle occipital gyrus (**Table S6, sFig 3**).

*Chronic Exclusive Neck/Shoulder Pain vs. Controls*

People reporting exclusive chronic neck/shoulder pain showed no significant differences in cortical or subcortical structure (**Table S6, sFig 3**).

*Chronic Exclusive Back Pain vs. Controls*

Compared to controls, people reporting chronic exclusive back pain showed lower surface area in the anterior insula, lateral occipito-temporal sulcus, and the pars triangularis (**Table S6, sFig 3**).

*Chronic Exclusive Abdominal Pain vs. Controls*

People reporting exclusive chronic abdominal pain showed no significant differences in cortical or subcortical structure (**Table S6, sFig 3**).

*Chronic Exclusive Hip Pain vs. Controls*

People reporting exclusive chronic hip pain had no significant differences in cortical or subcortical structure compared to controls (**Table S6, sFig 3**).

*Chronic Exclusive Knee Pain vs. Controls*

People reporting chronic exclusive knee pain had on average, greater cortical thickness in the superior parietal gyrus, and lower subcortical volume in the cerebellum and brainstem (**Table S6, sFig 3**).

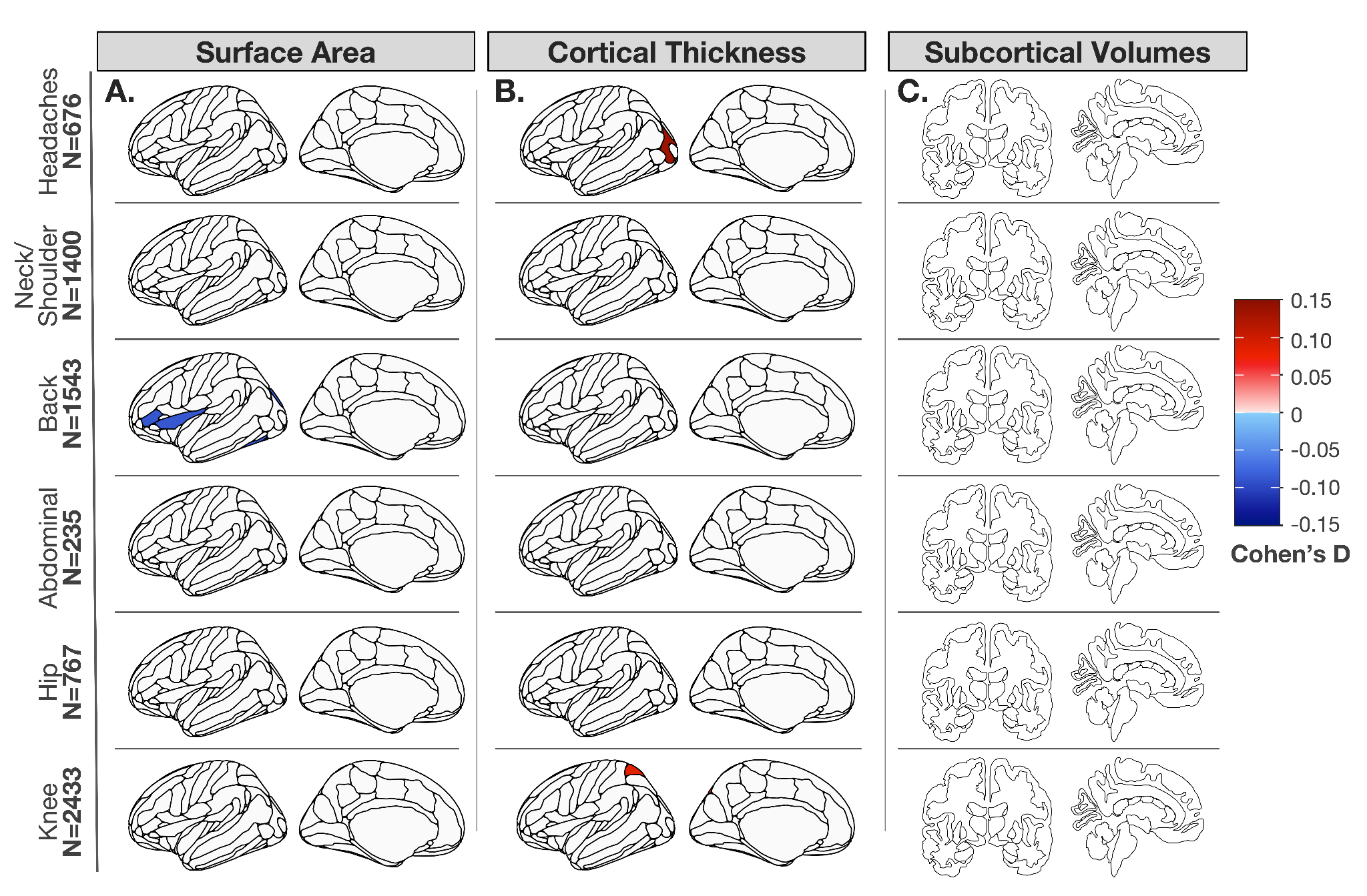

**sFigure 3:** Significant differences in brain structure in participants reporting chronic pain exclusively across different body regions compared to controls. Effect sizes (Cohen’s D) for surface area (A), cortical thickness (B), and subcortical volumes (C) of regions that are significant between groups.

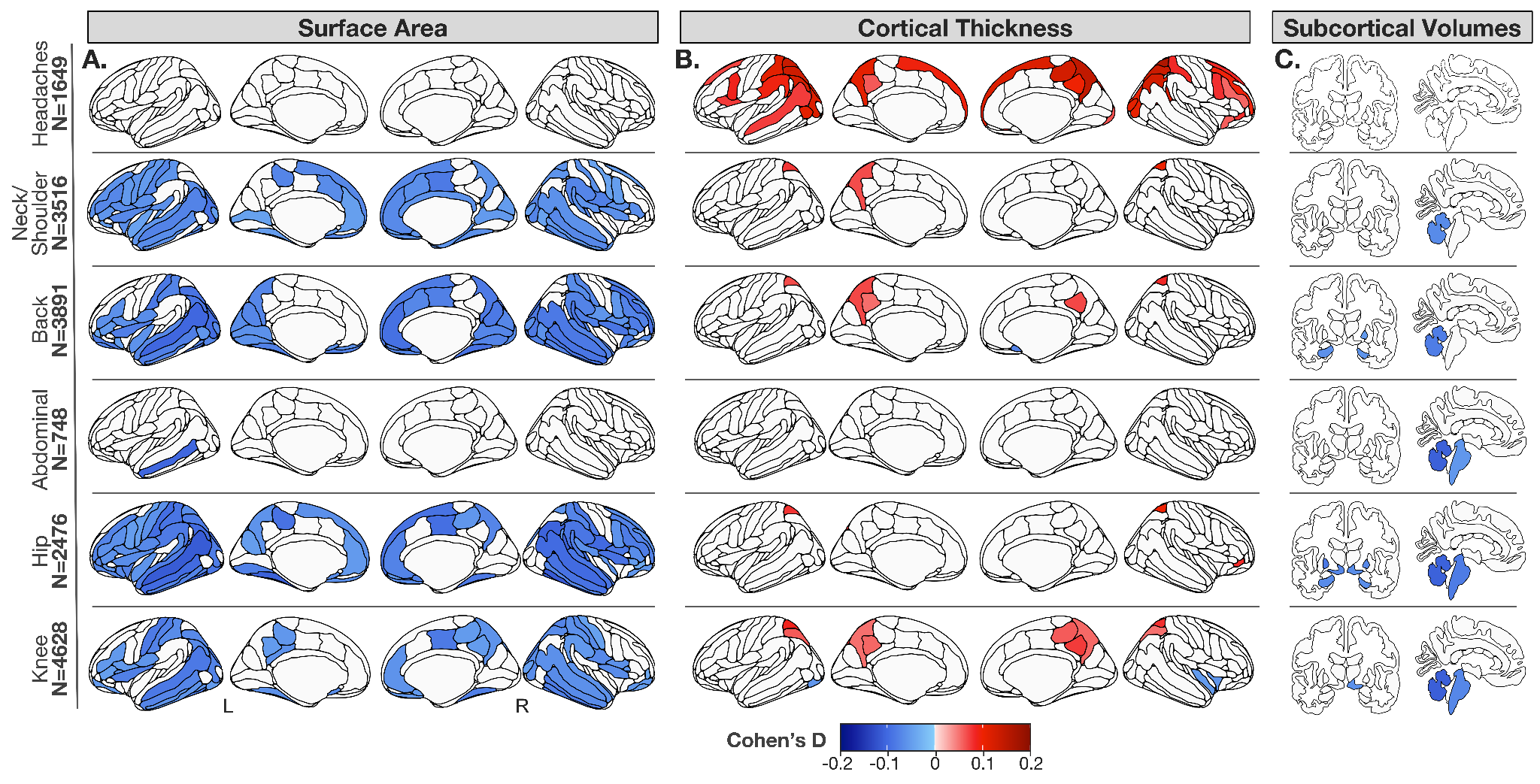

**sFigure 4:** Significant differences in brain structure in participants reporting chronic pain across different body regions compared to controls using a lateralized approach. Effect sizes (Cohen’s D) for surface area (A), cortical thickness (B), and subcortical volumes (C) of regions that are significant between groups.

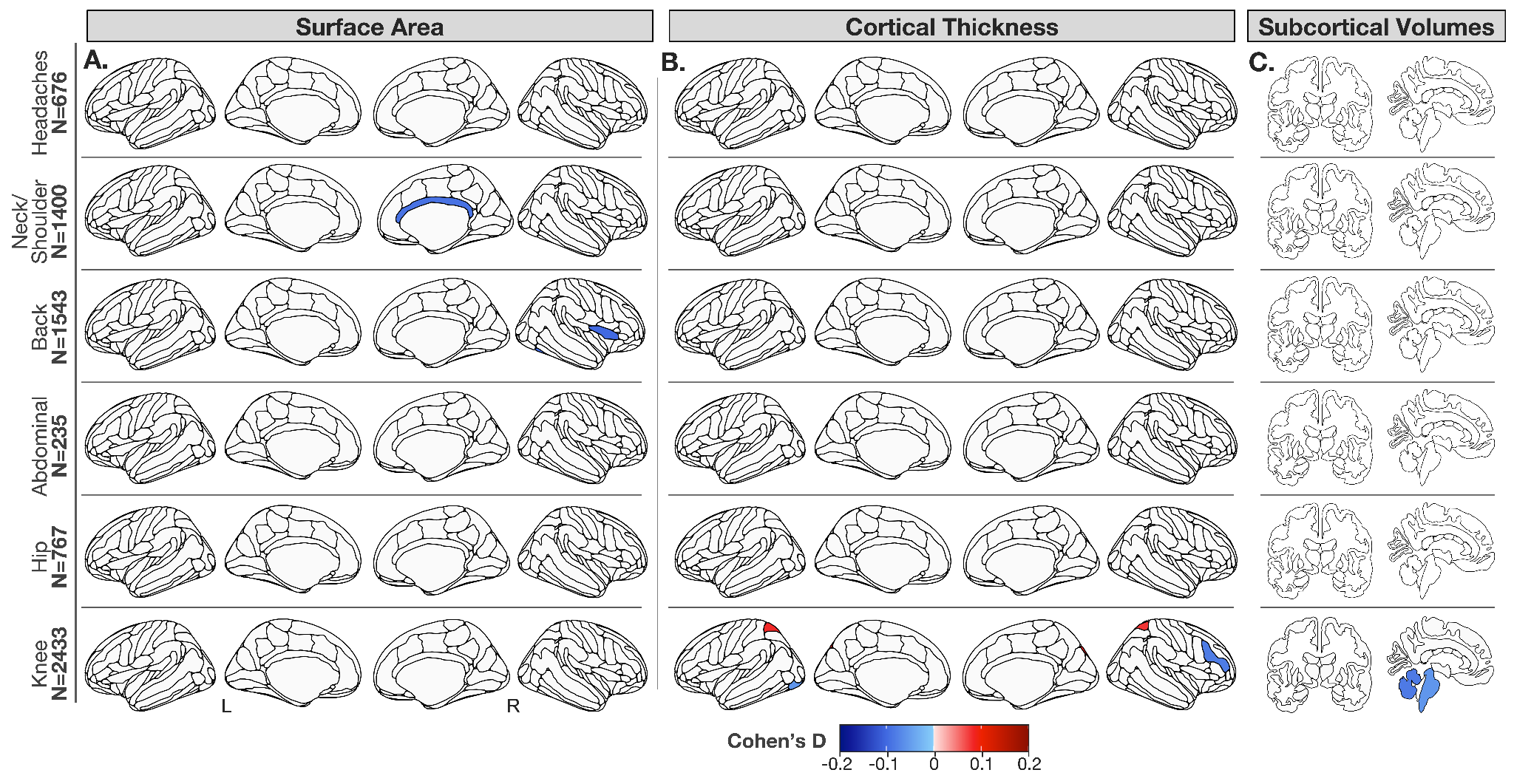

**sFigure 5:** Significant differences in brain structure in participants reporting chronic pain exclusively across different body regions compared to controls using a lateralized approach. Effect sizes (Cohen’s D) for surface area (A), cortical thickness (B), and subcortical volumes (C) of regions that are significant between groups.

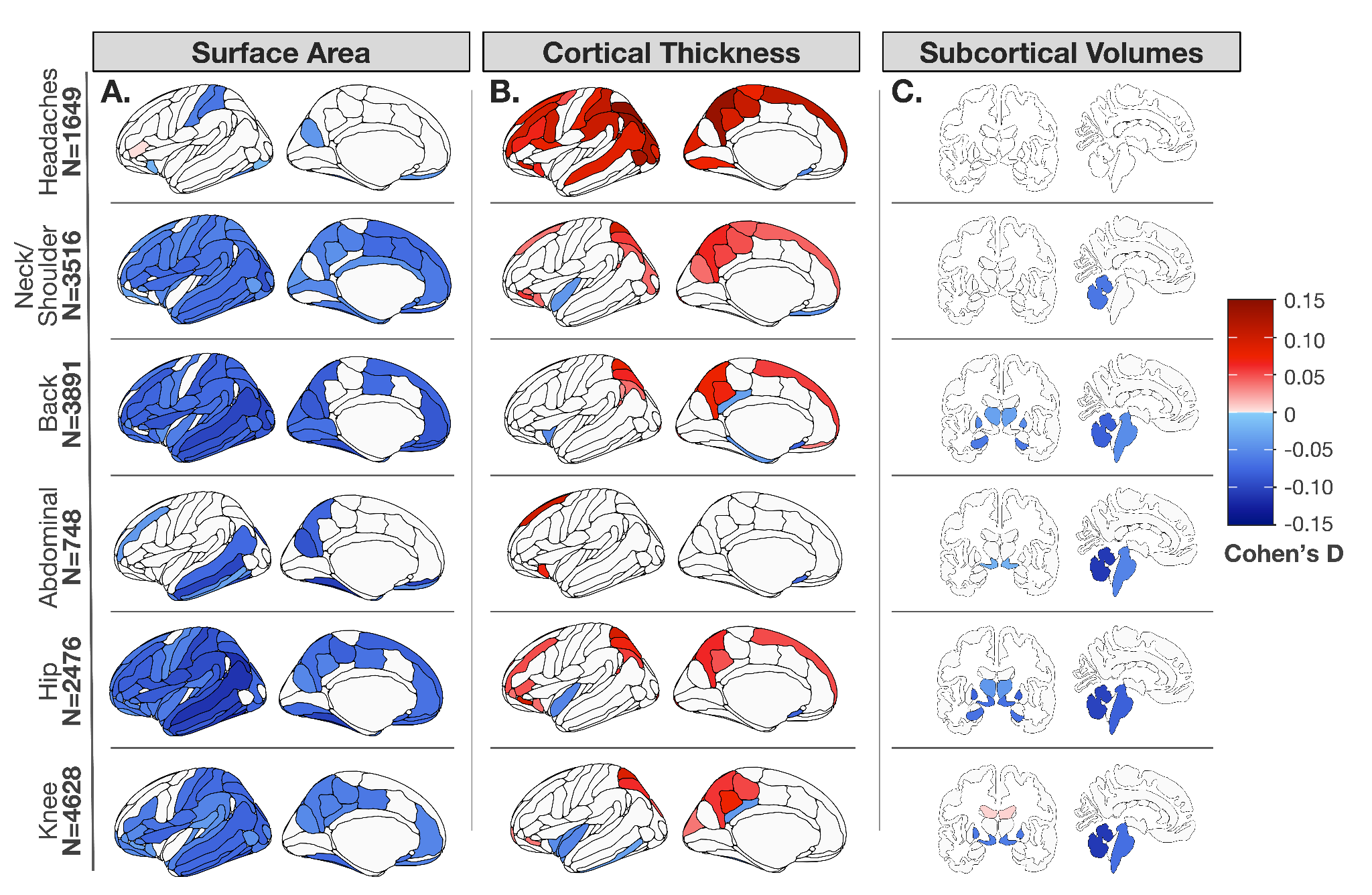

**sFigure 6**: Significant differences uncorrected for multiple comparisons in brain structure in participants reporting chronic pain across different body regions compared to controls. Effect sizes (Cohen’s D) for surface area (A), cortical thickness (B), and subcortical volumes (C) of regions that are significant between groups.

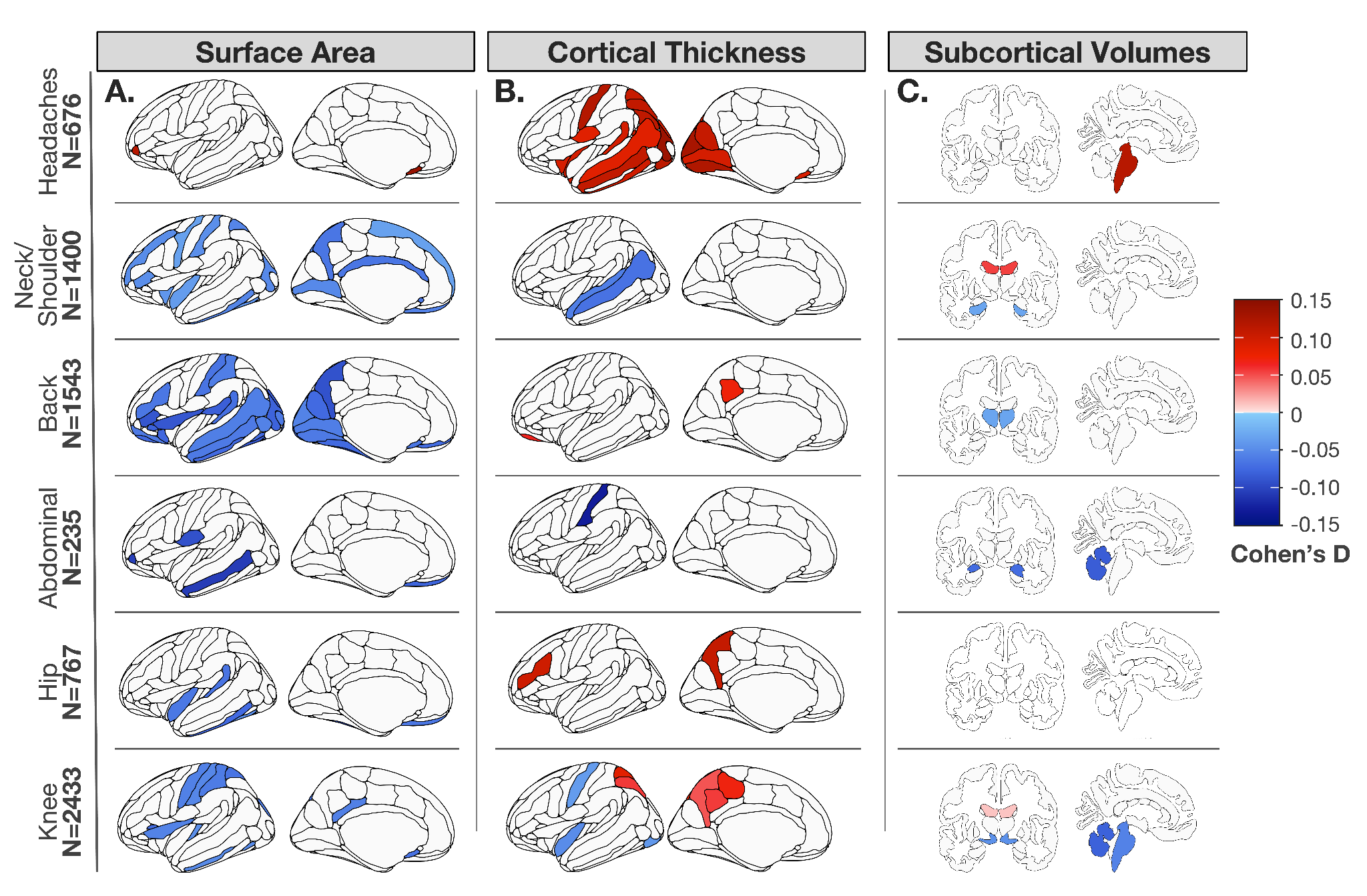

**sFigure 7**: Significant differences uncorrected for multiple comparisons in brain structure in participants reporting chronic pain exclusively across different body regions compared to controls. Effect sizes (Cohen’s D) for surface area (A), cortical thickness (B), and subcortical volumes (C) of regions that are significant between groups.
