## Supplementary material for "Brain Structural Differences in Adults Reporting Localized Chronic Pains Mediate Risk for Suicidal Behaviors": Table 1

**Table 1:** Demographics of participants with chronic pain matched to controls by age and sex. TDI: socioeconomic status via the Townsend deprivation index, eTIV: estimated total intracranial volume. IQR: interquartile range. The Wilcoxon rank sum and Pearson chi-squared tests were used on continuous and categorical variables, between CP and controls, respectively.

| **Characteristic** | **N** | **Chronic Pain, N = 11 298^1^** | **No Chronic Pain, N = 11 298^1^** | **p-value^2^** | **Cohen's D** | **Cramer's V** |
| --- | --- | --- | --- | --- | --- | --- |
| **Age** | 22 596 | 64 (58 – 70) | 64 (58 – 70) | 0.99 | 1.70e-04 |  |
| **Sex** | 22 596 |  |  | 1 |  | 0.00 |
| Female |  | 6 240 (55%) | 6 240 (55%) |  |  |  |
| Male |  | 5 058 (45%) | 5 058 (45%) |  |  |  |
| **TDI** | 19 198 | 2·63 (0·51 – 3·90) | 2·65 (0·59 – 3·93) | 0.379 | -0.0124 |  |
| Unknown |  | 2 190 | 1 208 |  |  |  |
| **eTIV mm^3^** | 22 596 | 1.42e+06 (1.33e+06 – 1.52e+06) | 1.42e+06 (1.33e+06 – 1.52e+06) | 0.00863 | -0.017 |  |
| **Anxiety** | 18 716 | 4 830 (54%) | 4 425 (45%) | 1.08e-38 |  | 0.095 |
| Unknown |  | 2 429 | 1 451 |  |  |  |
| **Depression** | 18 755 | 1 917 (22%) | 1 255 (13%) | 4.29e-61 |  | 0.12 |
| Unknown |  | 2 462 | 1 379 |  |  |  |
| ^1^Median (IQR); n (%) | | | | | | |
| ^2^Wilcoxon rank sum test; Pearson's Chi-squared test | | | | | | |
