## Supplementary material for "Brain Structural Differences in Adults Reporting Localized Chronic Pains Mediate Risk for Suicidal Behaviors": Table 2

**Table 2: Mediating Effect of Brain Regions on the Relationship Between Chronic Pain and Suicide**

| ***Brain Region*** | ***Pain Condition*** | ***B*** | ***95% CI*** | ***S*** | ***p*** |
| --- | --- | --- | --- | --- | --- |
| *Cerebellum Cortex Volume* | *Chronic Pain* | 1.61x10^-4^ | 5.05x10^-5^, 2.94x10^-4^ | 7.0x10^-4^ | 0.003 |
|  | *Neck/Shoulder Pain* | 1.54x10^-4^ | 3.49x10^-5^, 3.10x10^-4^ | 6.0x10^-4^ | 0.008 |
|  | *Back Pain* | 1.85x10^-4^ | 5.99x10^-5^, 3.43x10^-4^ | 7.0x10^-4^ | 8.0x10^-4^ |
|  | *Abdominal Pain* | 2.89x10^-4^ | 1.67x10^-5^, 6.28x10^-4^ | 6.0x10^-4^ | 0.03 |
|  | *Knee Pain* | 1.61x10^-4^ | 7.25x10^-5^, 3.37x10^-4^ | 6.0x10^-4^ | 0.03 |
| *Left Cerebellum Cortex Volume* | *Chronic Pain* | 1.67x10^-4^ | 5.99x10^-5^, 2.92x10^-4^ | 7.0x10^-4^ | 0.003 |
|  | *Abdominal Pain* | 3.02x10^-4^ | 6.13x10^-5^, 6.47x10^-4^ | 6.0x10^-4^ | 0.022 |
|  | *Knee Pain* | 1.70x10^-4^ | 2.90x10^-5^, 3.39x10^-4^ | 7.0x10^-4^ | 0.028 |
| *Right Cerebellum Cortex Volume* | *Chronic Pain* | 1.45x10^-4^ | 3.31x10^-5^, 2.75x10^-4^ | 6.0x10^-4^ | 0.009 |
|  | *Neck/Shoulder Pain* | 1.48x10^-4^ | 2.45x10^-5^, 3.45x10^-4^ | 5.0x10^-4^ | 0.01 |
|  | *Back Pain* | 1.70x10^-4^ | 4.72x10^-5^, 3.32x10^-4^ | 6.0x10^-4^ | 0.003 |
| *Right Superior Parietal Gyrus Thickness* | *Neck/Shoulder Pain* | 1.37x10^-4^ | 1.70x10^-5^, 2.96x10^-4^ | 5.0x10^-4^ | 0.023 |
| *Left Precuneus Thickness* | *Back Pain* | 1.30x10^-4^ | 2.80x10^-5^, 2.70x10^-4^ | 5.0x10^-4^ | 0.036 |

Abbreviations: *B:* Unstandardized indirect effect, 95% CI: 95% confidence interval (min, max), *S*: S-statistic, *p*: p-value
